# Chronic fatigue and post-exertional malaise in people living with long COVID

**DOI:** 10.1101/2021.06.11.21258564

**Authors:** Rosie Twomey, Jessica DeMars, Kelli Franklin, S. Nicole Culos-Reed, Jason Weatherald, James G. Wrightson

**Author notes:** **Corresponding author:** Rosie Twomey, Ph.D., @rosie_twomey. **Please cite as:** Twomey R, DeMars J, Franklin K, Culos-Reed SN, Weatherald J, Wrightson JG. Chronic fatigue and post-exertional malaise in people living with long COVID. *MedRχiv*. DOI: 10.1101/2021.06.11.21258564.

## Abstract

**Purpose:** People living with long COVID describe a high symptom burden, and a more detailed assessment of chronic fatigue and post-exertional malaise (PEM) may inform the development of rehabilitation recommendations. The aims of this study were to use validated questionnaires to measure the severity of fatigue and compare this with normative data and thresholds for clinical relevance in other diseases; measure and describe the impact of PEM; and assess symptoms of dysfunctional breathing, self-reported physical activity/sitting time, and health-related quality of life.

**Methods:** This was an observational study involving an online survey for adults living with long COVID (data collection from February-April, 2021) following a confirmed or suspected SARS-CoV-2 infection. Questionnaires included the Functional Assessment of Chronic Illness Therapy-Fatigue Scale (FACIT-F) and DePaul Symptom Questionnaire-Post-Exertional Malaise.

**Results:** After data cleaning, *n*=213 participants were included in the analysis. Participants primarily identified as women (85.5%), aged 40-59 (78.4%), who had been experiencing long COVID symptoms for ≥6 months (72.3%). The total FACIT-F score was 18±10 (where the score can range from 0-52, and a lower score indicates more severe fatigue), and 71.4% were experiencing chronic fatigue. Post-exertional symptom exacerbation affected most participants, and 58.7% met the scoring thresholds used in people living with myalgic encephalomyelitis/chronic fatigue syndrome. PEM occurred alongside a reduced capacity to work, be physically active, and function both physically and socially.

**Conclusion:** Long COVID is characterized by chronic fatigue that is clinically relevant and is at least as severe as fatigue in several other clinical conditions, including cancer. PEM appears to be a common and significant challenge for the majority of this patient group. Patients, researchers, and allied health professionals are seeking information on safe rehabilitation for people living with long COVID, particularly regarding exercise. Fatigue and post-exertional symptom exacerbation must be monitored and reported in studies involving interventions for people with long COVID.

## Introduction

Long COVID is a major public health concern. The scale of the COVID-19 pandemic means that even if a small proportion of people infected with SARS-CoV-2 have prolonged symptoms, this translates to millions of people worldwide [1]. Estimates suggest that 13.7% of people will continue to have symptoms 12 weeks after infection [2]. This is not only a phenomenon that affects hospitalized patients or those with comorbidities [3]. Even in individuals at low risk of COVID-19 mortality, chronic symptoms can be present and can co-occur with impairment in one or more organs [4]. Long COVID is a complex, heterogeneous condition that is defined based on an elapsed acute infectious period [5]. Although there is not complete consensus, long COVID has been defined as the presence of signs and symptoms that develop during or following an infection consistent with COVID-19 and continue for four weeks or longer [6,7]. Long COVID (alternatively called post-acute COVID-19 after 12 weeks) is characterized by persistent, debilitating and wide-ranging symptoms that vary between individuals but often include fatigue, shortness of breath, dry cough, cognitive impairment, palpitations, chest tightness, and dizziness [7,8]. Persistent (chronic) fatigue is consistently reported to be the most prevalent symptom of long COVID [3,8,9]. Chronic fatigue describes a distressing, persistent feeling of weariness, tiredness or exhaustion that is not alleviated by rest and is not proportional to recent activity levels. Chronic fatigue is a hallmark of multiple conditions, and is known to interfere with usual function and negatively impact quality of life [10,11].

Healthcare professionals who are living with long COVID describe its trajectory as unpredictable, episodic and having a relapse-remitting nature [12–14]. Reports of persistent fatigue alongside fluctuating symptoms that worsen unpredictably or in response to exertion have led to comparisons between long COVID and other post-viral conditions, including myalgic encephalomyelitis/chronic fatigue syndrome (ME/CFS) [5,6,15–17]. One of the hallmark symptoms of ME/CFS is post-exertional symptom exacerbation, also called post-exertional malaise (PEM) [18,19]. PEM is a worsening of symptoms and reduction in function after physical, cognitive, or emotional activity that would not have caused a problem before illness [18–20]. To date, the identification of PEM in people living with long COVID has been driven by one remarkable patient-led effort that asked participants to identify PEM based on a definition of PEM [8,9]. However, to date, preliminary studies exploring progressive exercise have not reported any measure of PEM at enrollment, nor reported monitoring tolerability or symptom exacerbation in response to exertion [21,22]. This may be because the presence of PEM in people living with long COVID has not been clearly identified using self-report tools that are validated in people with ME/CFS.

Several studies have focused on persistent symptoms, recovery or rehabilitation after hospital admission due to COVID-19 [3,23–26]. In contrast, few studies have included people who were not hospitalized (the majority of individuals affected by COVID-19) or did not have a laboratory diagnosis of COVID-19, but who have experienced an acute illness equivalent to the acute symptomatic presentation of COVID-19 and/or had a known exposure to the virus. There are several explanations for the lack of laboratory confirmation, including lack of access caused by limited polymerase chain reaction (PCR) testing capacity early in the pandemic; false-negative PCR tests (∼50% of tests during a 0–5-day incubation period, and ∼10% of cases after this point [27]); negative, i.e. inconclusive antibody or serology tests [28]; and the numerous disincentives to seeking or accessing testing that can disproportionately affect disadvantaged populations (such as stigma or potential loss of income) [29]. This issue has been consistently raised by patients who may be considered ineligible for long COVID health services and sickness benefits [30]. Therefore, it is important to include everyone with a confirmed or suspected SARS-CoV-2 infection when characterizing the symptom burden of long COVID.

The aim of this study was to perform a more detailed assessment of fatigue and PEM in people with long COVID to inform the development of physiotherapy/rehabilitation recommendations. In a sample of adults who identified as living with long COVID, the specific objectives of this study were to use validated questionnaires to (1) measure the severity of fatigue and compare this with normative data and thresholds for clinical relevance in other diseases; (2) assess PEM using screening methods recommended for use in people living with ME/CFS; and (3) describe symptoms of dysfunctional breathing, self-reported physical activity/sitting time, and health-related quality of life to compare with normative data, where available.

## Methods

### Study Design and Setting

This was an observational study involving an online survey hosted on Qualtrics, a web-based survey tool. The survey opened on February 11, 2021, and the final response was completed on April 25, 2021. The study was approved by the University of Calgary Conjoint Health Research Ethics Board (REB21-0159) and performed according to the Declaration of Helsinki, with the exception of preregistration. A STROBE checklist is available in Supplementary File 1 (S1). All supplementary files are available on the Open Science Framework (https://osf.io/dxu63/).

### Participants

The survey explicitly targeted people living with long COVID. Participants were presented with four eligibility criteria as follows:

1. You are an adult (aged ≥ 18 years)
2. You consider yourself to be experiencing long COVID. Other terms include “long hauler” or “post-COVID-19 syndrome.” Regardless of the name, you are eligible if you have long-term symptoms due to COVID-19, and your symptoms do not pre-date the confirmed or suspected infection with COVID-19.
3. You tested positive for COVID-19, or you strongly suspect that you were infected with COVID-19 but were either unable to access a COVID-19 test or tested negative (e.g., if you were tested early or late after exposure to the virus). Strong suspicion could be based on experiencing common COVID-19 symptoms during the acute phase of the virus (typically 2-14 days), having close contact with a confirmed case, and being linked with an outbreak.
4. It has been four weeks or longer since you received a positive COVID-19 test or since you first experienced symptoms of COVID-19.

The survey was delivered in English and required high-school-level reading comprehension, and to complete the survey, participants required access to a computer (desktop, laptop, or tablet) or smartphone. Therefore, non-English speakers and people without access to this technology were excluded from participating.

### Recruitment and Informed Consent

Participants were recruited from long COVID networks on social media (Twitter and Facebook). RT and JDM shared a recruitment slide (S2) with community leaders, patient advocates and patient support groups (where permission was granted) via these social networks. In addition, the authors shared the slide with their network of physiotherapy/rehabilitation professionals. A snowball recruitment strategy was used to allow patients and clinicians to identify other people living with long COVID. Study details such as the purpose, research team, the risks and benefits of participating, and contact details for more information were communicated on the first page of the survey using an implied consent approach. Participants were then presented with an agreement to participate, and consent was implied by a specific action (the decision to select “Yes, I consent”).

### The Online Survey

The web-based survey tool was set up to exclude identifiable information, including email addresses and Internet Protocol addresses. Participant names and contact details were not collected, and therefore responses could not be used to identify individuals. Participants were asked to select or describe their gender, select their age category, and self-report their country of residence. Participants were asked to select or describe diagnosed comorbidities that pre-dated COVID-19. Participants were asked to select the date of COVID-19 testing that resulted in a positive test (if applicable) and select or describe symptoms that accompanied the acute phase of the illness. Participants were also asked to select or describe their current ability to work (if applicable). The full survey is presented in S3. Participants were then presented with five questionnaires that were selected due to their psychometric properties, recommended use, and low participant burden to complete.

#### Fatigue

The Functional Assessment of Chronic Illness Therapy Fatigue Scale (FACIT-F) [31] is a 13-item self-report questionnaire that was developed for use in people with cancer and is widely recommended for the measurement of fatigue severity and its impact [32]. This study does not validate the use of the FACIT-F in people living with long COVID, and we do not claim that the FACIT-F items capture all aspects of fatigue in this population. However, the FACIT-F does have clinical utility for measuring fatigue outside of oncology, including in people living with HIV, lupus, rheumatoid arthritis, psoriatic arthritis, anemia, COPD, Parkinson’s disease and post-stroke [33–39]. In people living with and beyond cancer, the FACIT-F has a cut-point (<34, where the range is 0-52 and a lower score indicates more severe fatigue) that was designed to operationalize diagnostic fatigue criteria [40,41]. Although we do not claim that this is valid to “diagnose” fatigue in people with long COVID, we report the proportion of people who score below 34 as a crude comparison of clinical relevance and severity.

#### Post-exertional malaise

Historically, there has been difficulty in defining and measuring PEM [18,19], but for the purposes of this study, PEM was used to denote concepts related to post-exertional symptom exacerbation [18]. PEM was measured using the DePaul Symptom Questionnaire-Post-Exertional Malaise (DSQ-PEM) [42,43]. The DSQ-PEM asks participants about symptoms over a 6-month timeframe. However, we stipulated in the questionnaire instructions that if symptoms had been present for less than six months, participants should consider symptoms since the acute phase of COVID-19 or a positive COVID-19 test. Step 1 involves scoring above a threshold for one or more of the first five items. A threshold score of 2-4 for frequency (half the time, most of the time, or all of the time) coupled with a score of 2-4 for severity (moderate, severe, or very severe) for the same item is indicative of PEM. This method has been recommended by the National Institute of Neurological Disorders and Stroke (NINDS; part of the National Institute of Health) Common Data Elements (CDE) PEM working group [44]. Secondly, we added to the above method using items in the additional supplementary questions that cover quick recovery, exercise exacerbation, and PEM duration and were designed to operationalize the CDE recommendations further [45]. In addition to step 1, items 7 or 8 must have an answer of “Yes,” and a response of ≥14 h is required for item 9. However, the use of the supplementary items in this way was intended for the discrimination of people with ME/CFS from other conditions [45], and this was not our aim. Here, we use it as a stricter method of identifying PEM, considering our use of the DSQ-PEM outside of the context of ME/CFS. The DSQ-PEM was not used to comment on whether any of the participants in the present study have ME/CFS because this would require a more comprehensive clinical evaluation involving differential diagnoses and identification of other core symptoms [46,47].

#### Breathing

The 25-item Self-Evaluation of Breathing Questionnaire (SEBQ) was used to measure breathing discomfort related to perceptions of air hunger and the work or effort of breathing [48,49]. No cut-point has been validated, but in line with a recent study, we used a strict threshold of >25 as an indicator of significant breathing discomfort [50].

#### Health-Related quality of life

Health-related quality of life was measured using a 36-item instrument for adults, the RAND 36-Item Short-Form Health Survey (SF-36) [51]. The SF-36 measures eight multi-item health concepts (physical functioning, role limitations due to physical health problems, role limitations due to personal or emotional problems, energy/fatigue, emotional wellbeing, social functioning, bodily pain, and general health perceptions). It also includes a single item that provides an indication of perceived change in health. Responses are recoded so that each item is scored from 0-100%, with higher scores defining a more favourable health state. This questionnaire is a generic health-related quality of life tool that is useful for comparing general and specific populations and the relative burden of a health condition [52].

#### Physical activity/inactivity

Self-reported physical activity and sitting time were measures using the 7-item International Physical Activity Questionnaire-Short Form (IPAQ-SF) [53]. Data were processed according to standardized protocols available at www.ipaq.ki.se. The IPAQ-SF measures the self-reported frequency and duration (min) of vigorous-intensity activity, moderate-intensity activity, and walking performed in bouts of ≥10 minutes in the past week. Total weekly minutes for these activities were calculated, converted to metabolic equivalents (MET) and expressed as MET-minutes per week [54,55]. A summary indicator was used to categorize physical activity as ‘high’ (either 3 days of vigorous-intensity activity plus an accumulation of ≥1500 MET-min/week, or ≥5 days of any combination of activity that resulted in ≥3000 MET-min/week), ‘moderate’ (either ≥3 days of vigorous-intensity activity of ≥20 min/day, ≥5 days of moderate-intensity activity or walking for ≥30 min/day, or ≥5 days of any combination of activity that resulted in ≥600 MET-min/week), or ‘low’ (no physical activity reported or activity below the criteria for ‘moderate’). The IPAQ-SF also measures the duration (minutes per day) of time spent sitting on a usual weekday. This includes time spent sitting at a desk, visiting friends, or reading, or sitting or lying down while watching television across various contexts, including work, home, or leisure [53]. This data was reported as minutes per day.

### Data Analysis

Our primary analysis is descriptive and comparative, and data are presented as mean ± standard deviation, median (interquartile range), or frequency (percentage). In our companion paper (*under preparation*), we provide a qualitative analysis of the rich information on the participant experience provided in the optional free-text comments, analyzed via thematic analysis. In the present analysis, we checked all free-text comments for any indication that the participant was unsure how to answer a question, that the questionnaire options were not applicable, or that questionnaire answer(s) should be considered against additional information that could invalidate the response. In exploratory analyses, we checked for differences between groups dichotomized as (i) participants who did and did not receive a laboratory diagnosis of COVID-19; (ii) participants who did and did not have one or more medical condition that pre-dated COVID-19 (recoded as yes, no); and (iii) participants who did and did not report PEM, based on scoring step 2. Differences in categorical variables between groups were tested using Chi-squared tests. Differences in questionnaire scores between groups were tested using Mann-Whitney U tests, and these *p*-values were corrected for multiple comparisons using the Holm-Bonferroni correction. The relationships between the FACIT-F score and SF-36 subscales and SEBQ score were examined using Spearman’s correlations, with an adjusted false discovery rate. The threshold to reject the null hypothesis of no difference between groups was set as *p*<0.05. Analysis was performed using Jamovi [56] and R [57]. Some participant information is presented here in aggregate but was removed from the open dataset to further ensure participant anonymity. The open (quantitative) dataset and analysis are available at https://osf.io/dxu63/.

## Results

A total of *n*=280 people implied their consent, and *n*=211 participants (75.4%) completed 100% of the survey. Two researchers (RT, JGW) independently inspected survey responses for evidence of poor data quality (careless responses) and previous history of ME/CFS or post-viral fatigue, and these records were excluded. Both researchers checked all records for non-differentiation in ratings, particularly where items are reversed (FACIT-F and SF-36) and conducted random checks on data consistency (equivalent responses for similar items both within and between questionnaires). For example, inconsistencies across questionnaires were checked for items related to fatigue/tiredness on the FACIF-F and SF-36, vigorous and moderate activity on the SF-36 and IPAQ-SF, activity limitations on the SF-36 and DSQ-PEM, and exercise on the DSQ-PEM and IPAQ-SF. Both researchers individually inspected all complete records where no optional comments were provided (8.2%) and all records that participants completed in <10 minutes (6.7%). Partially complete records were included in the analysis if at least 50% of the survey was complete (this was equivalent to the participant completing both the FACIT-F and the DSQ-PEM), at least one optional comment regarding the experience of long COVID was provided, and no careless responses could be identified (*n*=8). One partially complete and two complete records were excluded because the participant indicated that they were asymptomatic or experienced one symptom during the acute phase of a suspected infection, did not received a positive test and provided no free-text elaboration for the suspected infection. Data cleaning procedures are outlined in detail in S4. Following data cleaning, a total of *n*=213 participants were included in the analysis. Excluding records where the time taken to complete the survey was more than two hours (*n*=6, assumed to have left the survey open while taking a break) and partially complete records, participants completed the survey in 27.4 ± 17.4 minutes.

### Participants

Participant characteristics are presented in Table 1. Participants primarily identified as women (85.5%), most were aged 40-49 (32.9%), 30-39 (23.5%) or 50-59 (22.1%), most were from the UK (39.5%), Canada (35.2%) or the USA (16.0%) and the sample overwhelmingly identified as white (93.0%). A large proportion of participants had no medical conditions that pre-dated long COVID (46.5%), as described in Table 1.

**Table 1.** Participant characteristics

|  | *n* | % |
| --- | --- | --- |
| **Age Category** |  |  |
| 18-29 | 21 | 9.9 |
| 30-39 | 50 | 23.5 |
| 40-49 | 70 | 32.9 |
| 50-59 | 47 | 22.1 |
| 60-69 | 21 | 9.9 |
| 70-79 | 4 | 1.9 |
| **Gender\*** |  |  |
| Woman | 182 | 85.4 |
| Man | 27 | 12.7 |
| Non-binary | 3 | 1.4 |
| Prefer not to answer | 1 | 0.5 |
| **Population Group(s)\*** |  |  |
| White | 198 | 93.0 |
| Black or African American | 3 | 1.4 |
| Hispanic, Latino, Latina or Latinx | 3 | 1.4 |
| Asian or Asian American | 1 | 0.5 |
| British Indian | 1 | 0.5 |
| East Indian | 1 | 0.5 |
| Indigenous | 1 | 0.5 |
| Malagasy | 1 | 0.5 |
| Ashkenazi Jewish & White | 1 | 0.5 |
| Hispanic, Latino, Latina or Latinx & White | 1 | 0.5 |
| Middle Eastern or Northern African & White | 1 | 0.5 |
| Native & White | 1 | 0.5 |
| **Country of Residence\*** |  |  |
| United Kingdom | 84 | 39.5 |
| Canada | 75 | 35.2 |
| USA | 34 | 16.0 |
| Other Europe | 10 | 4.7 |
| Australia | 1 | 0.5 |
| Brazil | 1 | 0.5 |
| Nigeria | 1 | 0.5 |
| Qatar | 1 | 0.5 |
| Not reported | 7 | 3.3 |
| **Existing Medical Conditions\*** |  |  |
| None | 99 | 46.5 |
| Depression or Anxiety | 16 | 7.5 |
| Autoimmune Disorder | 10 | 4.7 |
| Hypothyroidism | 8 | 3.7 |
| Fibromyalgia | 7 | 3.3 |
| Migraines | 7 | 3.3 |
| Anaemia | 6 | 2.8 |
| Asthma (moderate to severe) | 6 | 2.8 |
| Hypertension | 6 | 2.8 |
| Inflammatory bowel disease | 4 | 1.9 |
| Endometriosis | 3 | 1.4 |

Numbers may not sum to 100.0% due to rounding. \*Selected or self-identified using free text.

### The acute phase of COVID-19

We listed 11 common COVID-19 symptoms (fever; dry cough; tiredness; shortness of breath; aches and pains; sore throat; diarrhea; conjunctivitis; headache; loss of taste and smell; a rash on skin or discoloration of fingers and toes) and participants could additionally self-identify those that were not listed. The most common free-text symptoms were chest tightness or pain, brain fog or other cognitive impairment, and dizziness or lightheadedness (Table 2). Participants self-identified many other symptoms, and we report those that were present in more than 5% of the sample in Table 2; of these, participants identified a median of 7 (6-9) symptoms. Over half of participants were not tested/did not receive a positive test for COVID-19 (*n*=127, 59.6%). This is congruent with the proportion of patients with symptom onset early in the pandemic (April 2020 or earlier; 49.3% of the current sample) when people were less like to get tested. However, of those *n*=86 participants whose COVID-19 was confirmed through laboratory testing, some did have access to a test/tested positive early in the pandemic (April 2020 or earlier; 26.7%) but most tested positive from Sept-Dec 2020 (59.3%), when daily cases in Europe and North America increased. Few of the participants in this study had been hospitalized (*n*=7, 8.1%).

**Table 2.** The acute and chronic experience after confirmed or suspected infection with COVID-19

|  | *n* | % |
| --- | --- | --- |
| **Acute Symptoms\*** |  |  |
| Shortness of Breath | 202 | 94.8 |
| Aches & pains | 171 | 80.3 |
| Headache | 170 | 79.8 |
| Tiredness / Fatigue | 164 | 77.0 |
| Dry cough | 135 | 63.4 |
| A rash on skin or discolouration of fingers and toe | 129 | 60.6 |
| Sore throat | 128 | 60.1 |
| Fever | 118 | 55.4 |
| Diarrhea | 82 | 38.5 |
| Loss of taste and smell | 37 | 17.4 |
| Chest tightness or pain | 31 | 14.6 |
| Conjunctivitis | 27 | 12.7 |
| Brain fog / confusion / cognitive impairment | 27 | 12.7 |
| Dizziness / light headedness | 23 | 10.8 |
| Rapid heart rate/ / tachycardia | 17 | 8.0 |
| Heart palpitations | 16 | 7.5 |
| Loss of appetite / weight loss | 13 | 6.1 |
| Nausea / vomiting | 12 | 5.6 |
| Sinus congestion / pressure | 11 | 5.2 |
| **Months experiencing long COVID symptoms** |  |  |
| 1-2 months | 20 | 9.4 |
| 3-5 months | 39 | 18.3 |
| 6-9 months | 30 | 14.1 |
| 10+ months | 124 | 58.2 |
| **Is long COVID preventing or limiting your ability to work?\*** |  |  |
| Yes - preventing my return to work / unable to work | 90 | 42.3 |
| Yes - able to work but usual capacity is limited / reduced hours | 89 | 41.8 |
| No or N/A - retired, not employed, or a stay-at-home parent | 22 | 10.3 |
| No - able to work as usual | 11 | 5.2 |
| Not reported | 1 | 0.5 |
| **Positive Test** |  |  |
| Yes | 86 | 40.4 |
| No | 127 | 59.6 |

Numbers may not sum to 100.0% due to rounding. \*Selected or self-identified using free text.

### Long COVID

The majority of the participants had been experiencing long COVID symptoms for more than six months (72.3%; Table 2). One-third (*n*=71) of participants indicated that they were not receiving support from their medical/healthcare team for long COVID symptoms. A large subset of participants indicated that long COVID was preventing their return to work or that they were unable to work (42.3%). A second large subset indicated that long COVID had reduced their capacity to work or reduced the hours they were able to work (41.8%). Only 5.2% of the participants were able to work as usual. The question on employment was not applicable for 10.3% of participants, though several elaborated on reduced capacity in other contexts (e.g., home and family roles).

### Fatigue (FACIT-F)

The total FACIT-F score was 18 ± 10 (where the score can range from 0-52, and a lower score indicates more severe fatigue). As a reference, means and SDs for other clinical conditions are presented in Table 3.

**Table 3.** FACIT-F scores in a range of other populations for comparison with long COVID.

| Disease or Clinical condition | FACIT-F* | *n* | Age* | %Female* | Study |
| --- | --- | --- | --- | --- | --- |
| Long COVID | $18 \pm 10$ | 213 | Table 1 | 85 | - |
| General population | $44 \pm 9$ | 1010 | $46 \pm 17$ | 52 | [58] |
| Cancer & anemia | $24 \pm 13$ | 2292 | $63 \pm 13$ | 65 | [58] |
| Chronic cancer-related fatigue | $27 \pm 7$ | 51 | $54 \pm 11$ | 65 | [59] |
| Human immunodeficiency virus | $34 \pm 13$ | 51 | $40 \pm 7$ | 12 | [33] |
| Rheumatoid arthritis | $29 \pm 11$ | 631 | 56<sup>‡</sup> | 79 | [36] |
| Psoriatic arthritis | $36 \pm 12$ | 135 | $52 \pm 13$ | 42 | [34] |
| Iron deficiency anemia | $24 \pm 12$ | 608 | $45 \pm 14$ | 89 | [38] |
| Chronic obstructive pulmonary disease | $42 \pm 9$ | 564 | $68 \pm 10$ | 68 | [60] |
| Parkinson's disease | $34 \pm 10$ | 118 | $64 \pm 10$ | 46 | [39] |
| Chronic immune thrombocytopenia | $36 \pm 12$ | 207 | 50<sup>‡</sup> | 67 | [61] |
| Stroke | $38 \pm 10$ | 51 | $63 \pm 14$ | 51 | [33] |

\*Values have been rounded for presentation. <sup>‡</sup>=median

More than 90% of the sample were below the cut-off for clinically relevant fatigue in people living with and beyond cancer (Table 4), and 71.4% were experiencing chronic fatigue, based on this cut-point and symptoms persisting for ≥3 months.

**Table 4.**
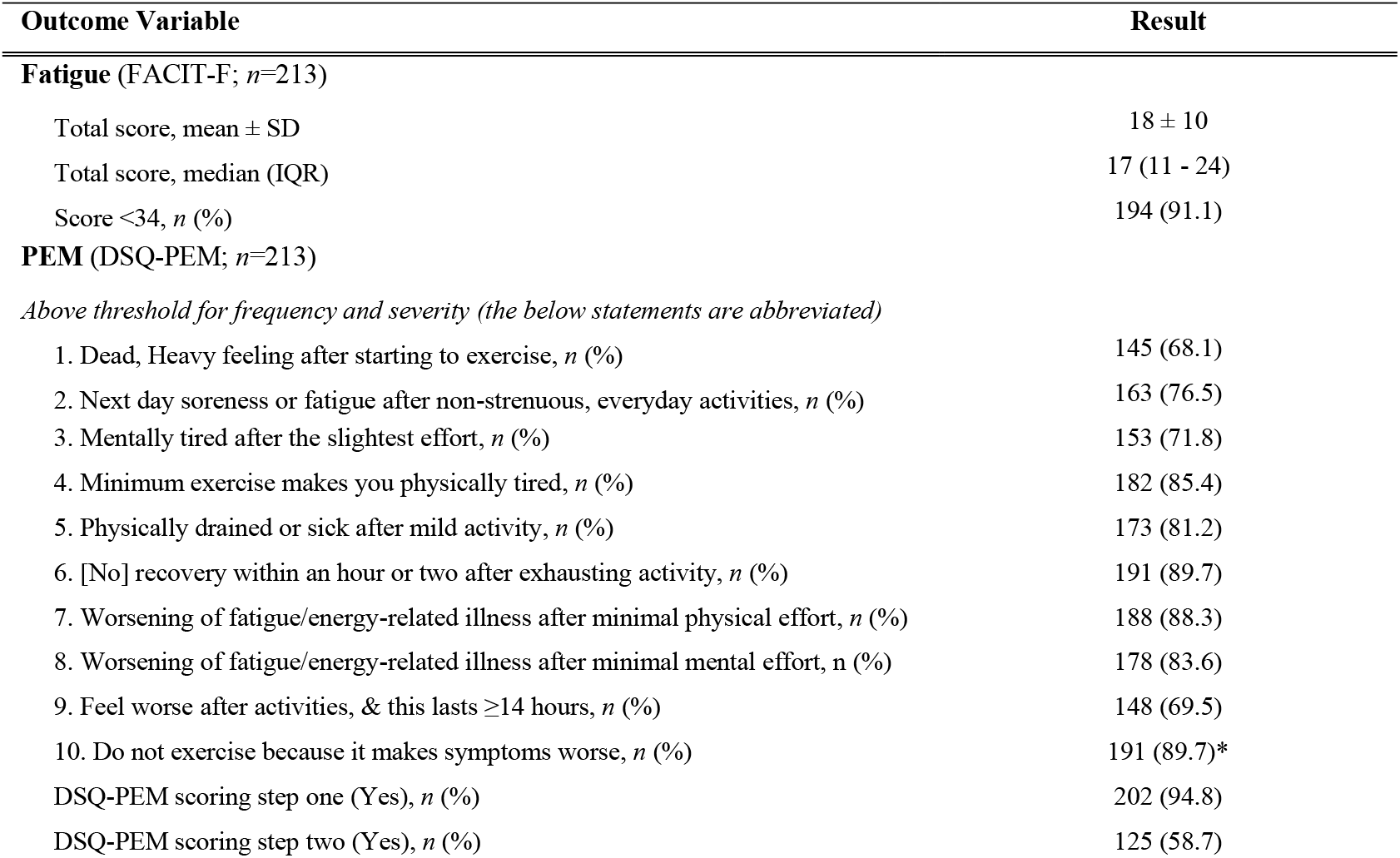
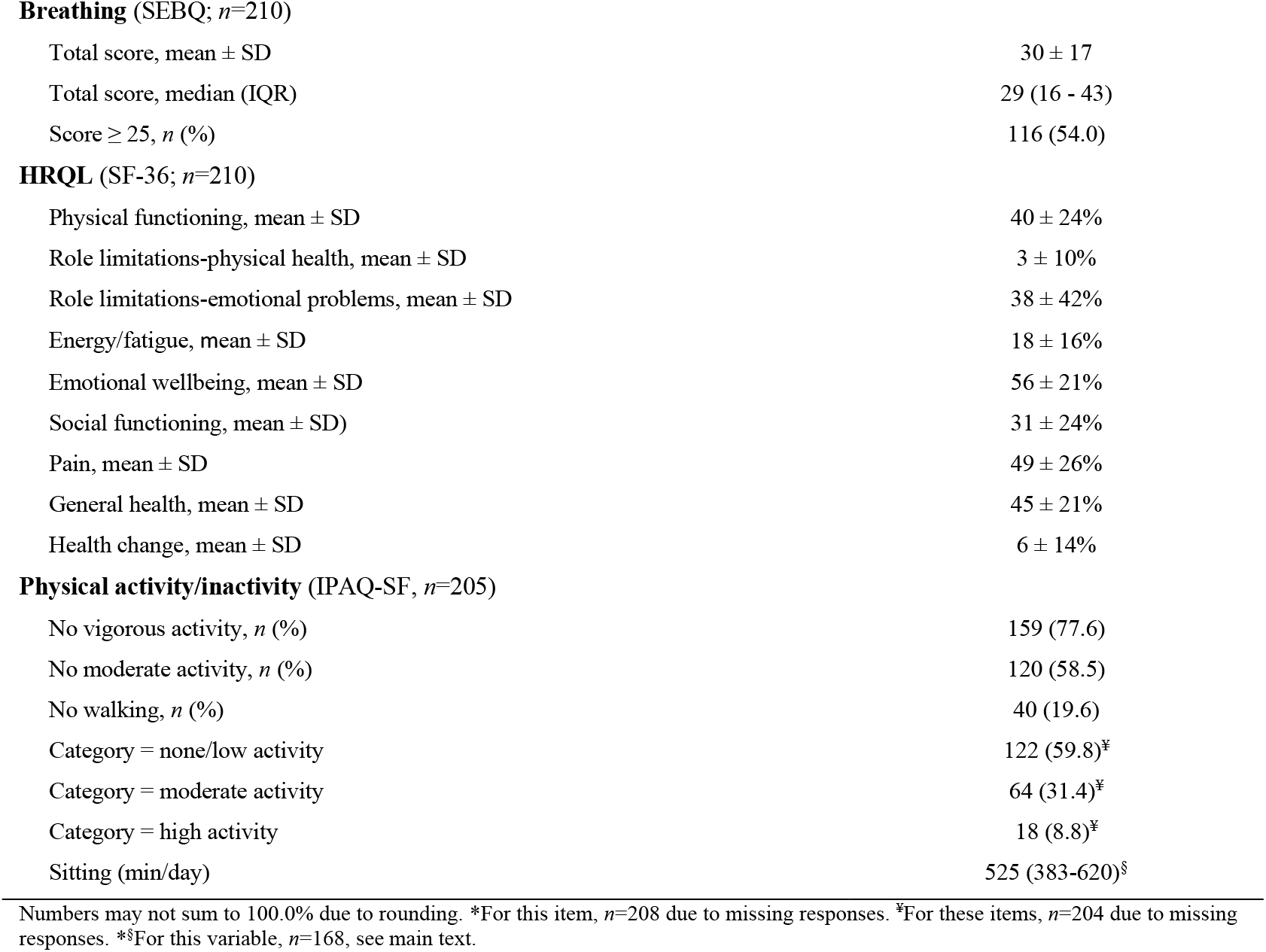
Patient-reported outcome measures

### Post-exertional malaise (DSQ-PEM)

In step 1, participants rated the frequency and severity of items 1-5, abbreviated in Table 4. The threshold for each item was a frequency ranging from half the time to all the time, combined with a severity of moderate to very severe. The proportion of participants meeting this threshold for these items ranged from 68.1% (*Dead, heavy feeling after starting to exercise*) to 85.4% (*Minimum exercise makes you physically tired*). Overall, 94.8% met the threshold for at least one of the first five items. However, only 4.7% met the threshold for *only* one of the first five items. Rather, nearly half (46.9%) of participants met the threshold for *all* of the first five items. In step 2, which incorporates the supplementary items, 58.7% of participants in this sample met the scoring thresholds used in people with ME/CFS.

### Breathing (SEBQ)

The total SEBQ score was 30 ± 17 (where the score can range from 0-75, and a higher score indicates more severe symptoms). In a sample of 180 participants from the general population, the mean ± SD was reported as 15.5 ± 11.5 [48]. Furthermore, 54.0% of the sample had a score >25, indicating significant breathing discomfort.

### Health-related quality of life (SF-36)

Summary data for SF-36 subscales can be found in Table 4. The health concepts that were most impacted by long COVID were role limitations due to physical health problems (3 ± 10%, where all scores range from 0-100% and a higher percentage indicates better health) and energy/fatigue (18 ± 16%). The health change item asks participants, “Compared to one year ago, how would you rate your health in general now?” Here, the score was also strikingly low (6 ± 14%), with 79.5% answering “Much worse now than one year ago” and 18.6% answering “Somewhat worse now than one year ago.” Nine participants (4.3%) left free-text comments to express uncertainty about whether to answer questions 33-36 (which all contribute to the general health subscale) based on their current health or health prior to COVID-19. Removing these participant’s responses due to this uncertainty did not change the descriptive statistics for the general health subscale, so they remain in our analysis. As visualized in Figure 1, HRQL was severely impaired in people living in long COVID, in comparison to normative data from the general population [62], people living with rheumatoid arthritis (RA) [63], and chronic obstructive pulmonary disease (COPD) [64]

**Figure 1.**
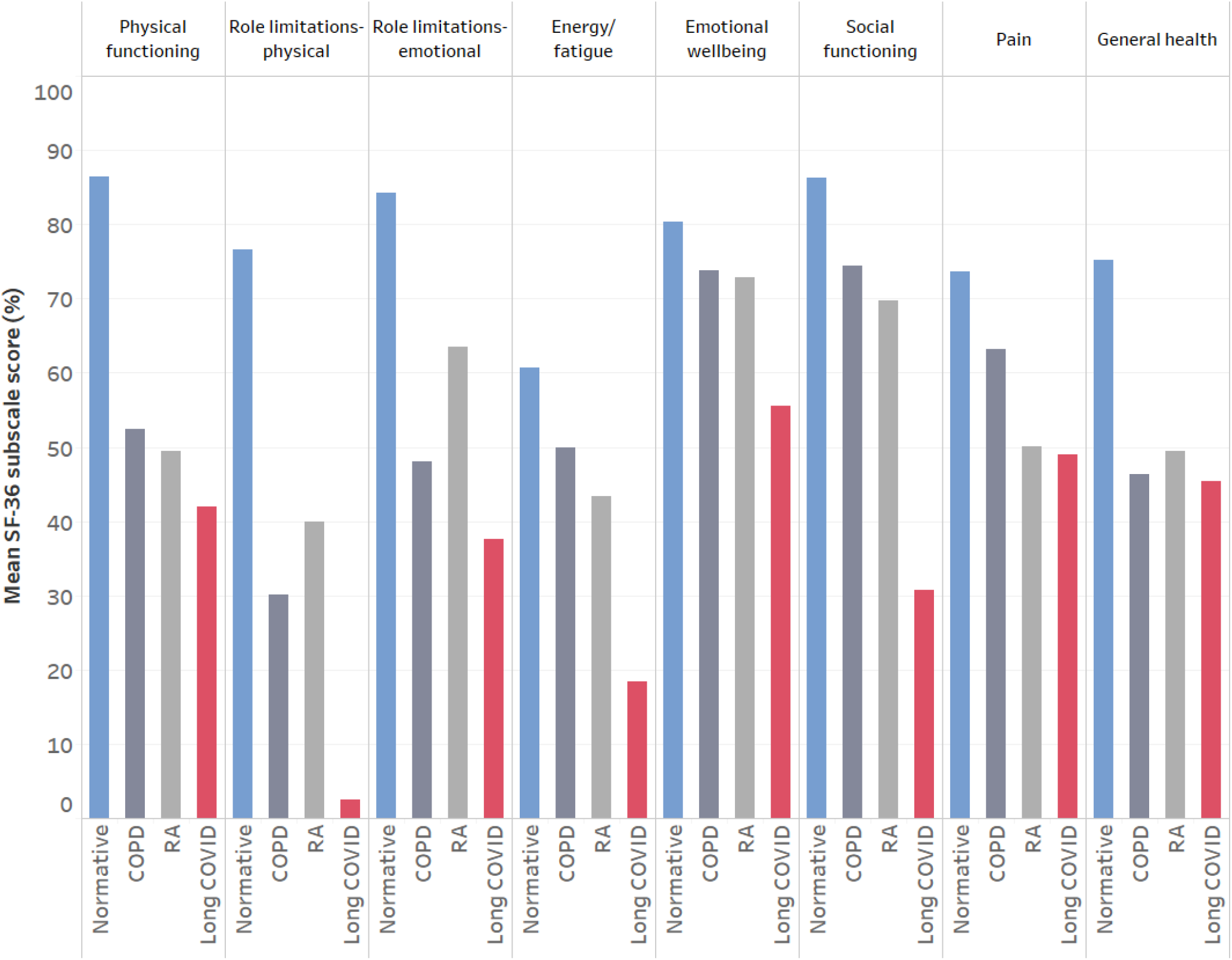
A visualization of the impact of long COVID on health-related quality of life measured using eight SF-36 subscales. Mean scores from the present study are presented alongside data from the general population (normative) [62], rheumatoid arthritis (RA) [63], and chronic obstructive pulmonary disease (COPD) [64].

### Physical activity/inactivity (IPAQ-SF)

Most participants reported no vigorous physical activity (77.6%, Table 4). In the remaining subset of participants (*n*=46) who did report vigorous physical activity, this took place on 2 (1-3) days per week, for 60 (30-65) minutes (median and IQR). However, most of these participants indicated that rather than those vigorous activities suggested by the IPAQ-SF (e.g., heavy lifting, digging, aerobics, or fast bicycling), they answered for typical daily activities, which were now rated as vigorous. This should be considered against the description of vigorous activities in the IPAQ-SF as “activities that take hard physical effort and make you breathe much harder than normal.” Over half of the participants reported no moderate physical activity (58.5%; note that the IPAQ-SF excludes walking from this category). In the remaining subset of participants (*n*=83) who did report moderate physical activity, this took place on 2 (1-4) days per week, for 50 (20-90) min, and participants made similar comments about the perceived effort of daily activities. Most participants reported some walking of at least 10 min duration (80.4%), and this took place on 4 (2-7) days per week, for 30 (20-60) min. Total physical activity over the past week was equivalent to 503 (99-1361) MET-minutes. Physical activity levels were categorized as none/low in 59.8%, moderate in 31.4%, and high in 8.8% of the sample (Table 4). Because the vigorous and moderate activity descriptors may not be valid in this population, later exploratory analysis uses only the walking variable as a measure of physical activity (because it is not based on perceived physical effort or breathing harder).

There were *n*=8 missing responses for time spent sitting for unknown reasons, though presumably because some participants found the wording unclear for this item. During data inspection, we noticed that some participants answered in total hours for five weekdays or for the full week rather than estimated a single weekday (and one participant left a comment to this effect). We also noticed that some participants included sleep in their estimate of sitting time (and one participant left a comment to this effect). Rather than devise a rule for correcting this data, we removed all responses >24 hours (*n*=18) and all responses between 19-24 hours (to remove data where sleep was included in the estimate; *n*=11). Therefore, the estimate of sitting time was in *n*=169 participants, who reported 525 (383-620) min [or 8.8 (6.4-10.3) hours] of time spent sitting per day on weekdays. However, this may be a poor surrogate for daily inactivity because *n*=10 of these participants left a comment to elaborate on spending time both sitting and lying down (or on the orthostatic challenges of sitting versus lying down. Because the IPAQ-PA specifically asks about time spent sitting as opposed to time spent awake but inactive, a median of 510 min may be an underestimate of sedentary time.

### Exploratory Statistical Analysis

#### Laboratory diagnosis

The only sample characteristic that was different between groups dichotomized based on laboratory diagnosis of COVID-19 was age (*χ*^*2*^=14.1; *p*=0.015), where participants aged 40-69 seemed to be less likely to have a laboratory confirmation (see S5 for contingency tables) in comparison to other age categories (Table 1). Participants who had a laboratory diagnosis did not have a higher occurrence of PEM (*χ*^*2*^=0.97; *p*=0.325), a higher fatigue severity (*U*=5138; *p*_*Holm*_=1.000; *r*=0.06) or more breathing discomfort (*U*=4938; *p*_*Holm*_=1.000; *r*=0.06). The only SF-36 subscale that differed between groups was general health (*U*=4044; *p*_*Holm*_*=*0.033; *r*=0.24), where people with laboratory confirmation rated their general health ∼10% better than those without. This was not due to differences in the proportion of people receiving support from their medical/healthcare team for long COVID symptoms (*χ*^*2*^=0.24; *p*=0.621).

#### Exploratory Relationships

More severe fatigue (FACIT-F score) was significantly associated with worse HRQL (all SF-36 subscales, S5), and more breathing discomfort (SEBQ; *rho*=-0.24 *p*_*fdr*_=0.002). The strongest correlations with the SF-36 subscale were with physical functioning (*rho*=0.65; *p*_*fdr*_=0.002; Figure 2), social functioning (*rho*=0.56; *p*_*fdr*_=0.002; Figure 2), pain (*rho*=0.40; *p*_*fdr*_=0.002; Figure 2) and health change (*rho*=0.33; *p*_*fdr*_=0.002).

**Figure 2.**
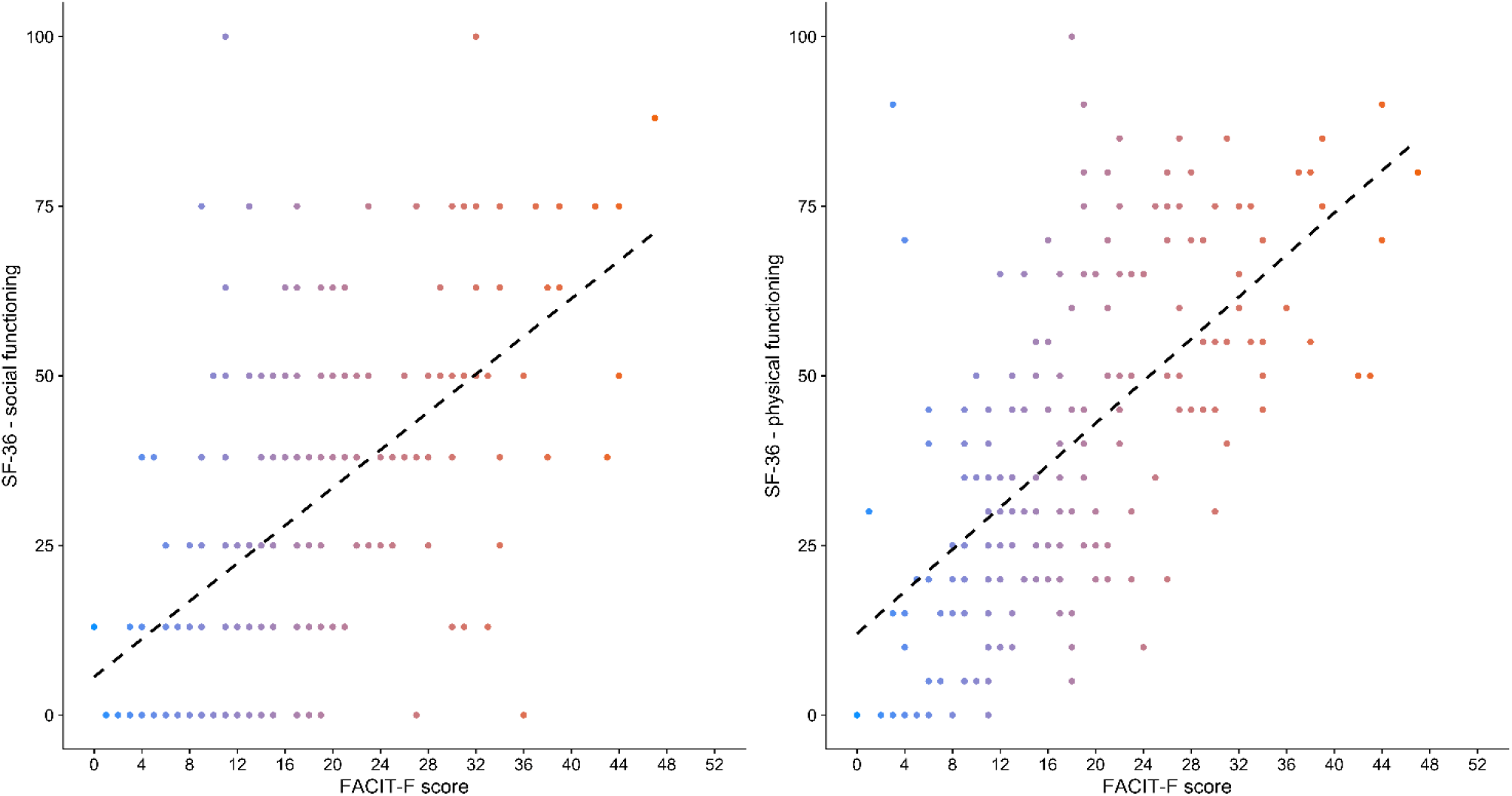
The relationship between fatigue (FACIT-F score, where a lower score represents more severe fatigue), social functioning (left panel; *rho*=0.56; *p*_*fdr*_=0.002) and physical functioning (right panel; *rho*=0.40; *p*_*fdr*_=0.002). Dashed lines are a graphical representation of Spearman’s correlation.

#### Medical/health conditions that pre-dated COVID-19

The only sample characteristic that was different between groups dichotomized based on medical/health conditions that pre-dated COVID-19 was support for long COVID symptoms (*χ*^*2*^=5.44; *p*=0.020); more participants with comorbidity reported receiving support from their medical/healthcare team. Participants with a comorbidity did not report more severe fatigue (*U*=4487; *p*_*Holm*_=0.080; *r*=0.20), but did report more breathing discomfort (*U*=3997; *p*_*Holm*_=0.011; *r*=0.26). Presence of any comorbidity was reflected in ∼10% lower physical functioning (*U*=3624; *p*_*Holm*_=0.011 *r*=0.34) and general health (*U*=3712; *p*_*Holm*_=0.011; *r*=0.32) subscales only (S5). Participants with comorbidity had a higher occurrence of PEM compared to those with none (64.9 vs. 51.5%; χ^2^=3.92; *p*=0.048).

#### Post-exertional malaise

Groups dichotomized based on the presence of PEM differed based on work status/limitations (*χ*^*2*^=13.0; *p*=0.023) and physical activity category (*χ*^*2*^=15.1; *p*<0.001). A large proportion (71.1%) of participants who reported that long COVID was limiting their usual capacity to work were experiencing PEM. Similarly, a large proportion (68.9%) of participants who reported no/low physical activity were experiencing PEM. Participants with PEM reported more severe fatigue (*U*=3888; *p*_*Holm*_=0.011; *r*=0.29; Figure 3), reduced physical functioning (*U*=3809; *p*_*Holm*_=0.011; *r*=0.29), reduced social functioning (*U*=3731; *p*_*Holm*_=0.011; *r*=0.30), and worse health compared to one year ago (*U*=4479; *p*_*Holm*_=0.028; *r*=0.16). There were no other differences in questionnaires (S5). In total, 95.2% of people with PEM scored <34 on the FACIT-F, compared to 85.2% of those without PEM (above the step 2 scoring threshold). Because comorbidities increase the prevalence of PEM by ∼13%, we repeated all of the above analyses for participants reporting no comorbidities (*n*=99) as a robustness check, and none of the above findings were altered.

**Figure 3.**
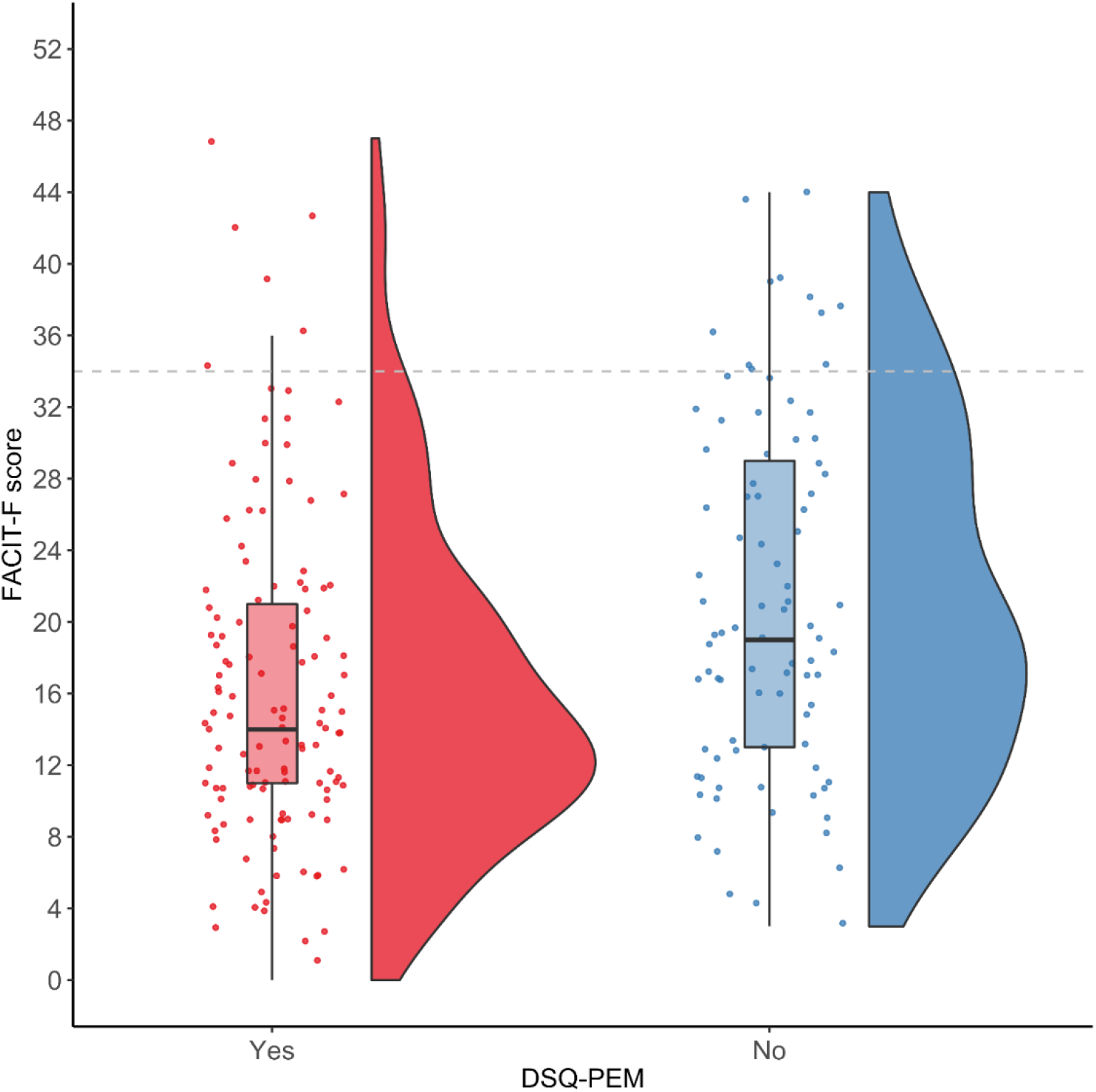
Raincloud plot [65] showing fatigue (FACIT-F score, where a lower score represents more severe fatigue) in people with post-exertional malaise, identified using step 2 scoring criteria for the DSQ-PEM questionnaire. Participants with PEM reported more severe fatigue (*p*_*Holm*_=0.011). As a crude marker of severity, the dashed line shows the threshold score of clinical relevance in oncology; recommended for the diagnosis of cancer-related fatigue.

## Discussion

In adults who identified as living with long COVID, our main findings were that (i) the overwhelming majority were living with chronic fatigue that was clinically relevant and appeared at least as severe as fatigue in several other clinical conditions, (ii) the impact on HRQL was substantial, despite a relatively young sample where almost half have no comorbidities, and (iii) the overwhelming majority were living with some level of post-exertional symptom exacerbation (or PEM), and many meet the threshold criteria for PEM using a self-report tool validated in people with ME/CFS. In this sample, people experiencing worsening of symptoms with exertion reported a reduced capacity to work and reduced physical and social functioning. Many participants were also living with breathing discomfort (air hunger and increased sensations of breathing effort), were unable to be physically active, had role limitations due to physical health problems, and rated their health as much worse compared to one year ago. Overall, symptom burden was not higher in people who received laboratory confirmation of COVID-19 compared to those with only an acute illness that was reasonably attributable to infection (in line with other data [66]), and comorbidities alone do not explain long COVID symptoms.

Chronic fatigue is difficult for patients to articulate and easy for others to dismiss [67,68]. Our data offer insight into the severity of the most common symptom, based on comparison with several other clinical conditions where the FACIT-F has been validated (Table 3), and comparison with a clinical cut-point recommended for the diagnosis of fatigue in people with cancer [40]. Fatigue is not only extremely common in long COVID, but for many, its severity and persistence are life-altering. Our exploratory analyses are in line with data from other populations on the associations between fatigue, reduced function, increased disability, and reduced HRQL [11,69]. The measurement of fatigue should be considered, and validation of a fatigue scale (such as the FACIT-F, which has demonstrated clinical utility in several populations) in people with long COVID is a priority. In addition, the use of visual analogue scales to measure response to intervention can be difficult to interpret if the measure (e.g., momentary or state perceptions of fatigue, or worst or average fatigue over a specific recall period) is not reported [22]. In fact, measurement of momentary fatigue using simple rating scales, such as the Rating of Fatigue scale [70] (pending validation in long COVID), might be best suited to the frequent monitoring of symptom exacerbation that is required of future reports of experimental interventions.

It is not currently understood how to treat chronic fatigue in people living with long COVID. Evidence from other conditions may offer some insight while data is being collected in long COVID. For example, at least some people with chronic fatigue after cancer treatment can benefit from exercise [71–73]. The mechanisms for the improvement in chronic cancer-related fatigue remain under investigation, but considering that the biological effects of exercise are multiple and interacting, reversal of deconditioning is unlikely to be the only pathway [74]. In contrast, exercise therapy is not a route to recovery for everyone with chronic cancer-related fatigue [71,75], and in people with ME/CFS, exercise (and other types of exertion) can cause serious setbacks and deterioration in function [20]. PEM is not caused by general deconditioning: PEM rarely occurs outside of the context of ME/CFS [75] and is associated with impairments measured during a two-day cardiopulmonary exercise test protocol that are not present in sedentary controls [76]. We found that a large proportion of people living with long COVID are experiencing PEM, and this corroborates many testimonials from patients/healthcare professionals with long COVID describing “relapses” after return to work and exercise [6,77]. Based on these findings, symptom exacerbation must be considered in rehabilitation and exercise interventions for people living with long COVID. In people with PEM, an activity plan needs to be carefully designed based on individual presentation with input from each patient [20]. The DSQ-PEM might be useful as a screening measure and to facilitate a discussion with patients about post-exertional symptom exacerbation. PEM will not be an issue for everyone with chronic fatigue, and the presence of PEM is not sufficient for an ME/CFS diagnosis. However, the presence of chronic fatigue and PEM that persists after COVID-19, alongside a substantial reduction in the ability to engage in pre-illness activities, unrefreshing sleep, cognitive impairment or orthostatic intolerance, should lead to a comprehensive assessment to exclude or diagnose ME/CFS [46,47], because this will help patients access appropriate care.

Considering PEM, beneficial interventions might first ensure symptom stabilization, with a long-term goal of improved function (for example, return to roles, daily activities or work [78]) and HRQL. In ME/CFS, pacing is a self-management strategy for activity that helps minimize severe symptom exacerbation [79]. Improvements may be aided by careful tailoring, pacing, prioritization, and modest goal setting [80]. More than half of our sample reported breathlessness and other respiratory symptoms. In the absence or in excess of the magnitude of physiological respiratory or cardiac disease, long COVID may involve chronic changes in breathing patterns that result in this breathing discomfort [81]. Respiratory physiotherapy and breathing retraining may be helpful for people with breathing discomfort, considering improvements in symptoms in people living with postural orthostatic tachycardia syndrome and asthma [81–84]. Currently, there is limited information about whether exercise is beneficial for people living with long COVID, especially considering the heterogeneous range of symptoms. While seeking insight from studies in other clinical conditions is valuable, the methodological/reporting inadequacies of rehabilitation literature [73,85] should not be repeated in studies of long COVID. While exercise is likely to be beneficial for some, there are many unknowns, including whether all patients with persistent symptoms should undergo screening for respiratory and cardiac complications before beginning exercise; whether exercise rehabilitation needs to be medically supervised; what level of tailoring is required; what frequency, intensity, duration and type of exercise can be recommended; and the trajectory of recovery for people living with long COVID. These uncertainties, and the fact that little data exists to date, mean treatment may require a multidisciplinary care pathway [86,87]. Limited reporting of interventions with overinterpretations regarding safety [21] do not serve the long COVID research, allied health professional, nor patient community. It is essential that future clinical trials (including pilot/feasibility studies) report modifications, symptom exacerbation and other adverse events. More generally, transparent reporting using published guidelines and open-access repositories [88– 91] will ensure progress in optimizing care for people with long COVID. Furthermore, research may be more impactful and meaningful when it is deeply collaborative and involves patients as partners [92].

### Limitations

One of the main limitations of survey designs such as ours is selection bias. People living with long COVID who were experiencing fatigue, PEM or breathlessness may have been more inclined to participate than people living with long COVID who were not experiencing any of these specific symptoms. Many participants accessed the survey via social media and online support groups, and the extent to which our sample is representative of the wider population of people living with long COVID is unknown. Our data suggest that a large proportion of people living with long COVID experience chronic fatigue, PEM, persistent breathing discomfort, and reduced HRQL. Although we were primarily interested in these concepts, we acknowledge the constellation of signs and symptoms that make up the long COVID experience that were not assessed here, including symptoms related to cognitive impairment and orthostatic intolerance. Furthermore, our sample was composed of primarily white participants from North America or Europe, and our data may not be generalizable to other racial/ethnic groups or other world regions. We intended to use self-report to estimate moderate and vigorous physical activity levels alongside PEM. However, because some participants rated activities of daily living as moderate or vigorous physical activity, the data were difficult to interpret. Including actigraphy may be necessary in future studies requiring an estimate of physical activity in this population.

## Conclusion

In this sample, long COVID is characterized by reduced HRQL and chronic fatigue that is clinically relevant and is at least as severe as fatigue in several other clinical conditions, including cancer. PEM seems to be a common and significant challenge for the majority of this patient group and occurs alongside a reduced capacity to work, be physically active, and function both physically and socially. Patients, researchers and allied health professionals are seeking information on safe rehabilitation and the potential role of exercise for people living with long COVID. Because people with long COVID report setbacks and deterioration in function following overexertion, fatigue and post-exertional symptom exacerbation must be monitored and reported in studies involving interventions for people with long COVID.

## Data Availability

An anonymized dataset is available at: https://osf.io/dxu63/

## CRediT author statement

Rosie Twomey: Conceptualization, Methodology, Software, Formal analysis, Data curation, Writing - original draft, Project administration.

Jessica DeMars: Conceptualization, Methodology, Writing - review and editing.

Kelli Franklin: Patient partner (provided guidance based on the lived experience of long COVID), Writing - review and editing.

Jason Weatherald: Writing - review and editing.

S. Nicole Culos-Reed: Writing - review and editing.

James G. Wrightson: Conceptualization, Formal analysis, Visualization, Writing - review and editing.

## Availability of data and material (data transparency)

An anonymized dataset is available at: https://osf.io/dxu63/

## Funding

This study was not funded. However, RT is supported by the O’Brien Institute of Public Health and Ohlson Research Initiative, Cumming School of Medicine, University of Calgary. JGW is supported by the Hotchkiss Brain Institute and the Cumming School of Medicine, University of Calgary. NCR is funded by the Canadian Cancer Society, Canadian Institutes of Health Research, Alberta Cancer Foundation, and University of Calgary funding support. JW is supported by the Libin Cardiovascular Institute at the University of Calgary, Heart & Stroke Foundation of Canada, and Canadian Institutes of Health Research.

## Conflicts of interest/Competing interests

JDM is a physiotherapist and owner of Breath Well Physio (Alberta, Canada) and has been treating people living with long COVID in private practice since July 2020. JDM and RT have piloted a free virtual program (Breath, Speak, Pace) for people living with long COVID in Alberta, Canada, funded by the Canadian Lung Association. This program will be continuing in partnership with Synaptic Health (Registered Charity No. 830838280RR001). JDM delivered a paid course for rehabilitation professionals working with people with long COVID in April 2021. The authors have no other conflicts of interest to disclose.

## Acknowledgements

The authors wish to sincerely thank the people living with long COVID who took part in this study for sharing their experiences.

## Data and supplementary materials

https://osf.io/dxu63/

S1: STROBE checklist

S2: Recruitment slide

S3: Survey

S4: Data cleaning procedures

S5: Reduced (anonymized) dataset

## Notes

### Clinical Trial

N/A

### Author Declarations

The study was approved by the University of Calgary Conjoint Health Research Ethics Board(REB21-0159).

